# Ambulance and hospital use after a coal mine fire: Analysis of linked data

**DOI:** 10.1101/2025.06.24.25330233

**Authors:** Tyler J. Lane, Caroline X. Gao, Catherine L. Smith, Jillian F. Ikin, Rongbin Xu, Matthew T.C. Carroll, Emily Nehme, Natasha Kinsman, Thara Govindaraju, Michael J. Abramson, Karen Walker-Bone, Yuming Guo

**Author notes:** Corresponding author Tyler J. Lane School of Public Health and Preventive Medicine Monash University 553 St Kilda Road Melbourne VIC 3004 Australia e. t.

## Abstract

**Background:** The 2014 Hazelwood coal mine fire shrouded the town of Morwell in regional Victoria, Australia, in smoke for six weeks. Building on previous analyses, we explored linked survey and healthcare data to investigate longer-term effects of fire-related PM_2.5_ exposure on ambulance attendances, emergency department presentations, and hospital admissions.

**Methods:** The Hazelwood Health Study Adult Survey cohort data were combined with modelled air pollution data to estimate individual levels of fire-related PM_2.5_ exposure and linked with ambulance, emergency department, and hospital admission data up to 2022. Associations between fire-related PM_2.5_ and health service use were evaluated using a recurrent event survival model over the 8-year post-fire period and within time strata (<2.5 years, 2.5-5 years, >5 years) and by condition, sex, and age.

**Results:** There were no detectable effects of fire-related PM_2.5_ on overall health service use in the total 8-year post-fire period, although there were some for specific conditions. PM_2.5_ exposure was associated with increased ambulance attendances for respiratory conditions, which attenuated over time, and injuries, which were sustained over the follow-up period. PM_2.5_ was also associated with temporary increase in cardiovascular emergency presentations and reduced cardiovascular hospital admissions. The reductions in cardiovascular emergency presentations and hospital admissions were only detectable among men.

**Conclusions:** The short-lived effect of PM_2.5_ on respiratory hospital admissions aligned with previous findings on the mine fire’s effects on respiratory health. The increase in injury-related ambulance attendances also aligns with prior findings and might be attributable to neurological damage from fire-related PM_2.5_ exposure. Reductions in cardiovascular health service use were new and likely attributable to immortal time bias due to previously-identified rises in cardiovascular death in the first months after the fire.

## 1 Introduction

On 9 February 2014, a bushfire ignited an open-cut brown coal mine in regional Victoria, Australia, shrouding the nearby town of Morwell in smoke and ash for six weeks (1). Daily mean fire-related particulate matter <2.5µm (PM_2.5_) was estimated to have reached 1022µg/m^3^ (2), nearly 7 times the Victorian Environmental Protection Authority’s 150µg/m^3^ threshold for “extremely poor” air quality (3).

Inhaled PM_2.5_ is linked to numerous health ailments including cardiopulmonary disorders, diabetes, kidney disease, neurological conditions like Parkinson’s and dementia, and cancer (4–8). Compared to typical sources of ambient PM_2.5_ in residential environments such as car exhaust and industry, the effects are likely worse when originating from landscape fires.

This could be partially due to the release of finer particles that can penetrate deep into the lungs, and from there, into the bloodstream and throughout the body (9–11). However, at the time of the mine fire, there was limited evidence about the health effects of smoky conditions lasting for weeks (12), which hampered the State Government’s response (1).

Due to this gap in the evidence, as well as community concerns, the Victorian Department of Health established the Hazelwood Health Study to evaluate the mine fire’s health effects (1).

Globally, exposure to smoke from landscape fires is associated with increased emergency presentations and hospital admissions, particularly for respiratory and cardiovascular conditions (13). Similarly, the Hazelwood Health Study has found numerous associations between smoke exposure and health service use. Analysis of population-wide, deidentified health service data indicated that ambulance attendances, emergency presentations, and hospital admissions increased during the mine fire, particularly for respiratory and mental health conditions (14–16). To better understand the mine fire’s effects, we analysed health service use data that were linked to a cohort survey containing rich, individual-level data on fire-related PM_2.5_ exposure and confounders (17). Prior analyses of these linked data indicated there were increased risks of ambulance attendances, emergency presentations, and hospital admissions in the 5 years after the fire, particularly for cardiopulmonary conditions (18–20). Here, we used an update linkage with 8 years of follow-up to address the following questions:

1. Was PM_2.5_ from the 2014 Hazelwood coal mine fire associated with long-term changes in ambulance attendances, emergency department presentations, or hospital admissions?
2. Did changes vary over time or by condition type, sex, and age group?

## 2 Methods

### 2.1 Data

The Hazelwood Health Study Adult Survey was administered to residents of Morwell (n = 3096), which neighboured the mine and experienced the most smoke exposure during the mine fire, and the minimally exposed control site Sale (n = 960), from May 2016 to February 2017, approximately 2-3 years post-fire. Of these 2,725 agreed to have their responses linked to administrative health datasets. Potential participants were identified through the Victorian Electoral Commission roll based on individuals who were 18 years or older and living in either Morwell or socioeconomically comparable areas of Sale at the start of the mine fire in February 2014 (17).

We linked Hazelwood Health Study’s Adult Survey cohort data to three health service datasets: Ambulance Victoria’s electronic patient care records for ambulance attendances (21), the Victorian Emergency Minimum Dataset (VEMD) for emergency presentations (22), and the Victorian Admitted Episodes Dataset (VAED) for hospital admissions (23) up to 30 June 2022. In addition, the data were linked to the National Death Index (24) to censor participants who died during follow-up. To categorise ambulance attendances by condition, we relied the final primary assessment, which reflected the most accurate evaluation made at the scene. In emergency and hospital admission data, we relied on primary diagnosis codes and used principal diagnosis in sensitivity analyses. The VEMD permits up to three diagnosis codes, with the first also representing the principal diagnosis. The VAED allows for up to 40 primary diagnoses, capturing conditions or underlying diseases that were present at the time of admission relevant to the episode of care. The first listed primary diagnosis is recorded as the principal diagnosis, which is the condition determined to be the main reason for admission after assessment. Codes and categories are listed in Table S1.

#### 2.1.1 Exposures

Due to insufficient air quality monitoring early in the fire, it was necessary to model fire-related PM_2.5_ concentrations (2). The estimates were generated through emissions and chemical-transport modelling, which factored in existing air monitoring, local weather conditions, coal combustion data. PM_2.5_ exposure was expressed as the daily average over the course of the mine fire in units of 10µg/m^3^ (2). These were then combined with participants’ time-location diaries that documented their whereabouts during the mine fire (17).

#### 2.1.2 Outcomes

Outcomes are time-to-event health service uses since the end of the mine fire. Health service use consisted of hospital admissions, emergency presentations, and ambulance attendances. Cause-specific hospital admissions and emergency presentations were determined using ICD-10-AM codes for principal diagnosis and categorised as cardiovascular, respiratory, mental health and injury conditions.

#### 2.1.3 Confounders

Confounders included number of health service uses pre-fire, age at the time of the fire, sex, employment (working, unemployed, student/volunteer/other), education (year 12 or less, certificate/diploma/tertiary), marital status (married/de facto, unmarried), work-related exposure (coal mine/power station, other occupational exposures [i.e., occupations exposed to dust, fumes, smoke, gas, vapour, or mist], unexposed), smoking (current, former, non-smoker [<100 lifetime cigarettes]), Index of Relative Socioeconomic Advantage and Disadvantage (IRSAD) score from the 2011 Census based on Statistical Area at Level 1 (25), and self-reported cardiovascular conditions, mental health disorders, or asthma/chronic obstructive pulmonary disease (COPD) that had been diagnosed by a doctor before the fire.

### 2.2 Statistical analysis

For descriptive analyses, we grouped participants into the following four groups: low, medium, and high exposure based on tertiles of mean daily fire-related PM_2.5_ during the mine fire period, and Sale residents as a separate group. These were then used for comparison of crude differences in health service use and confounders using one-way Analysis of Variance (ANOVA) for continuous variables and chi-squared tests for categorical variables.

To determine PM_2.5_ exposure effects, we applied the Prentice, Williams, and Peterson counting process (PWP-CP) (26), a recurrent survival model, as used in previous Hazelwood Health Study analyses on health service use (18–20). The PWP-CP model is an extension of the Cox survival model for recurring events that allows the baseline hazard to vary between events. To adjust for within-subject correlation, we used a robust variance estimator.

Stratified analyses analysed time-to-event outcomes within condition types and health services. In addition to evaluating post-fire changes in health service use over the entire follow-up period, analyses were time-stratified into the <2.5 years post-fire, 2.5-5 years post-fire, and >5 years post-fire, to account for time-varying effects observed in previous studies (19,27–29). All models adjusted for potential confounders. To stabilise estimates, health service uses were capped at the 95^th^ percentile within each analysis; e.g., overall analysis capped total health service uses at the 95^th^ percentile, while ambulance analysis capped health service use at the 95^th^ percentile of ambulance attendances.

Models were not evaluated when the number of events was low (<50 total events for primary analysis and <100 total events for time-stratified analysis) to avoid unstable estimates. Missing data were addressed using Multivariate Imputation by Chained Equations (MICE), with 20 imputed datasets and results pooled according to Rubin’s rules (30). All analyses were conducted using R in RStudio (31,32).

Due to substantial differences in healthcare service use between study sites observed in previous studies (20), main analyses excluded control site Sale; however, these data were included in sensitivity analyses. Other sensitivity analyses included using only principal diagnoses of hospital admissions and emergency presentations and capping the maximum number of health services in each analysis at 5.

### 2.3 Ethics

The Monash University Human Research Ethics Committee approved this analysis as part of the Hazelwood Adult Survey & Health Record Linkage Study (Project ID: 25680; previously CF15/872–2015000389 and 6066).

## 3 Results

### 3.1 Description of cohort

Out of 4,056 Adult Survey participants, 2,115 participants from Morwell and 610 participants from Sale consented to data linkage. As well as the expected differences between participants based on their mine fire-related PM_2.5_ exposure levels, Sale participants had higher proportions of TAFE/trade/tertiary education, fewer current and former smokers, and less occupational exposure overall, while unemployment was higher in Morwell, and PM_2.5_ exposure was associated with lower marriage rates.

Health service uses were generally low. However, there were crude differences between exposure groups. Ambulance attendances and hospital admissions were lowest in Sale, though the differences in ambulance attendances can only be inferred from the 75^th^ percentile as less than half of cohort members in all exposure groups were attended by an ambulance. There were no detectable differences by emergency presentations.

**Table 1.** Descriptive statistics, grouped by fire-related PM_2.5_ tertiles.

|  | Sale<br>(control)<br>(n=610) | Low<br>(n=705) | Morwell<br>Medium (n=718) | High<br>(n=692) | p value |
| --- | --- | --- | --- | --- | --- |
| **Daily mean PM<sub>2.5</sub> exposure** | 0 [0, 0] | 7 [5, 7] | 11 [10, 13] | 26 [19, 38] | <0.001 |
| **Female** | 345 (57%) | 384 (54%) | 380 (53%) | 369 (53%) | 0.581 |
| Missing | 0 | 0 | 2 | 0 |  |
| **Age** | 60 [47, 72] | 61 [50, 70] | 58 [47, 70] | 60 [49, 70] | 0.330 |
| **Highest educational qualification** |  |  |  |  | < 0.001 |
| Year 12 or below | 238 (39%) | 346 (49%) | 402 (57%) | 344 (50%) |  |
| TAFE†/trade cert/tertiary | 369 (61%) | 356 (51%) | 309 (43%) | 346 (50%) |  |
| Missing | 3 | 3 | 7 | 2 |  |
| **Employment** |  |  |  |  | <0.001 |
| Paid employment | 272 (45%) | 283 (40%) | 286 (40%) | 299 (43%) |  |
| Unemployed | 296 (49%) | 334 (48%) | 327 (46%) | 302 (44%) |  |
| Other (e.g., retired, volunteer, student) | 40 (6.6%) | 82 (12%) | 103 (14%) | 87 (13%) |  |
| Missing | 2 | 6 | 2 | 4 |  |
| **Married/de facto** | 394 (64.6%) | 466 (66%) | 439 (61%) | 382 (55%) | <0.001 |
| **Asthma or COPD before 2014** | 145 (24%) | 189 (27%) | 202 (28%) | 176 (26%) | 0.306 |
| Missing | 2 | 4 | 2 | 2 |  |
| **Cardiovascular diseases before 2014** | 131 (22%) | 127 (18%) | 156 (22%) | 150 (22%) | 0.244 |
| Missing | 3 | 6 | 5 | 4 |  |
| **Cancer before 2014** | 69 (11%) | 73 (10%) | 72 (10%) | 66 (9.6%) | 0.771 |
| Missing | 4 | 7 | 6 | 7 |  |
| **Smoking status** |  |  |  |  | 0.015 |
| Current smoker | 82 (14%) | 102 (15%) | 129 (18%) | 134 (19%) |  |
| Former smoker | 205 (34%) | 265 (38%) | 249 (35%) | 249 (36%) |  |
| Never | 318 (53%) | 331 (47%) | 337 (47%) | 306 (44%) |  |
| Missing | 5 | 7 | 3 | 3 |  |
| **Occupational exposure** |  |  |  |  | <0.001 |
| Coal mine/power station | 21 (3.4%) | 120 (17%) | 119 (17%) | 128 (18%) |  |
| Other occupational exposures | 205 (34%) | 184 (26%) | 198 (28%) | 186 (27%) |  |
| None | 384 (63%) | 401 (57%) | 401 (56%) | 378 (55%) |  |
| **Health service uses (per 10 person years)** |  |  |  |  |  |
| Ambulance attendances | 0 [0, 1] | 0 [0, 2] | 0 [0, 2] | 0 [0, 2] | <0.001 |
| Emergency presentations | 2 (1, 6) | 2 (1, 6) | 2 (1, 7) | 2 (1, 6) | 0.403 |
| Hospital admissions | 2 (1, 7) | 4 (1, 8) | 4 (1, 9) | 4 (1, 8) | <0.001 |

\*Statistical tests: Kruskal-Wallis for continuous variables, Fisher's exact test for dichotomous variables, and chi-squared for categorical variables. †TAFE: Technical and Further Education

### 3.2 Associations between fire-related PM_2.5_ exposure and health service use

Results from PWP-CP models are illustrated in Figure 1 and Table S2. We found no detectable associations between fire-related PM_2.5_ exposure and overall hospital admissions, emergency presentations, or ambulance attendances. Sensitivity analyses using participants from both Morwell and the control site Sale (see Table S3), principal diagnosis (see Table S4), and incorporating a 5-service cap (see Table S5), similarly found no detectable effects on overall health service use.

**Figure 1.**
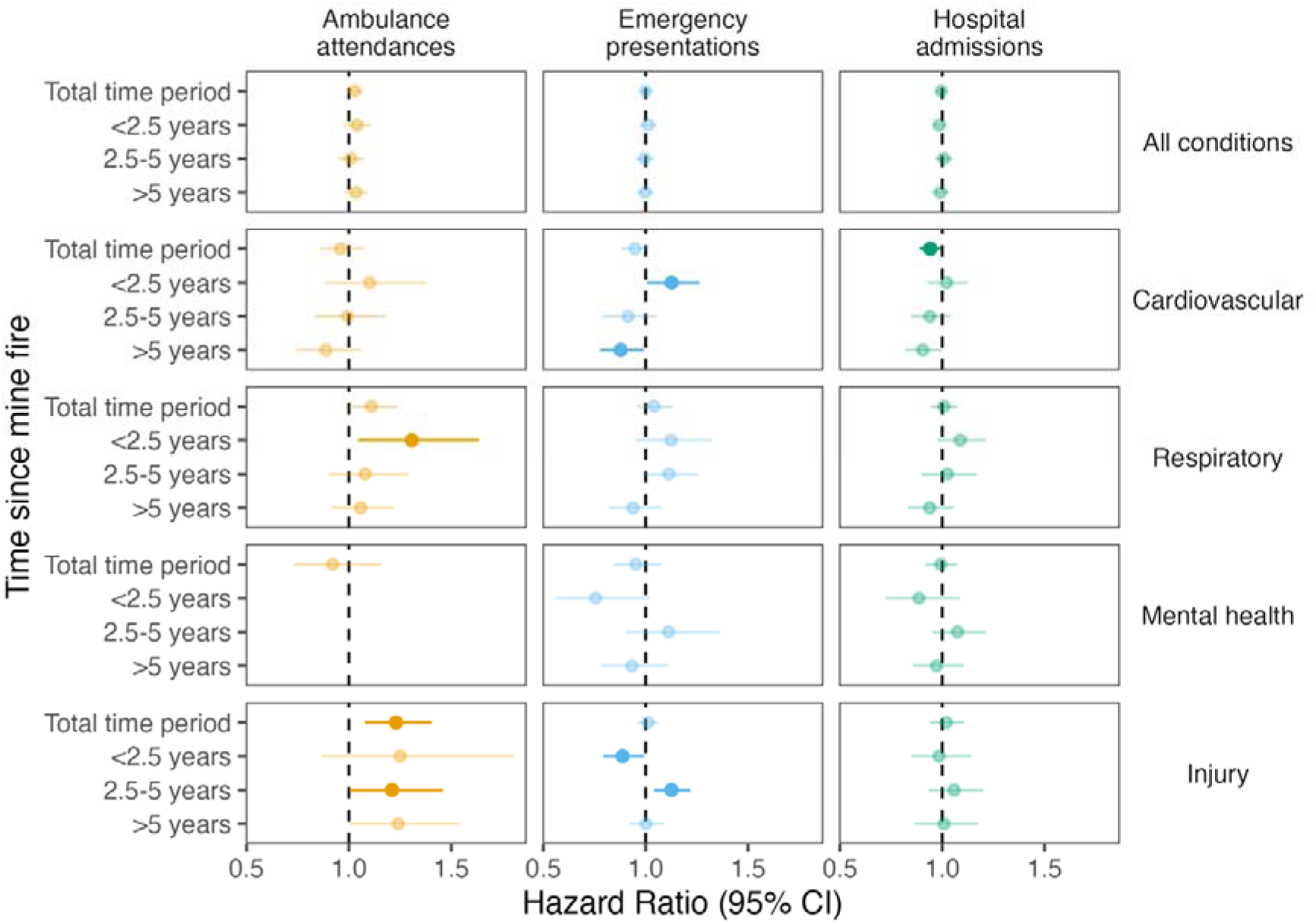
Changes in hospital admissions, emergency presentations, and ambulance attendances in association with a 10µg/m increase in daily mean mine fire-related PM_2.5_ exposure. Note: Hazard Ratios (HRs) were estimated from imputed PWP-CP models among Morwell participants, adjusting for confounding factors. Cause-specific hospitalisations and emergency presentations were obtained from all primary diagnoses. Faded points and CIs indicate non-significant (p > 0.05) estimates.

However, there were some detectable associations within condition groups. There was a 6% reduction in cardiovascular hospital admissions for every 10µg/m^3^ of daily average PM_2.5_ exposure (HR: 0.94; 95%CI: 0.89-1.00). The patterns over time were similar for ambulance attendances and emergency presentations, with an initial increase in services and later reduction, though these were only significant in emergency presentations.

There was limited evidence of an effect on health service use for respiratory conditions. There was 31% increase in respiratory ambulance attendances in the first 2.5 years post-fire (HR: 1.31; 95% CI: 1.04-1.64), which attenuated in the following years. While not reaching statistical significance in other health services, there was a similar pattern of initial rise and later fall for respiratory conditions in hospital attendances and emergency presentations.

Ambulance attendances for injuries increased 23% (95% CI: 1.08-1.40%) per 10µg/m^3^ of fire-related PM_2.5_ exposure over the total time period, with evidence that the effect persisted at each stratified time period. Emergency presentations for injuries were initially lower (<2.5 years HR: 0.89, 95% CI: 0.79-0.99) then higher (2.5-5 years HR: 1.13, 95% CI: 1.04-1.22) before attenuating to null afterwards (HR: 1.00; 95%CI: 0.92-1.09). There was no detectable association between fire-related PM_2.5_ and injury-related hospital admissions. Nor were there detectable effects on mental health service use.

### 3.3 Changes in health service use by sex and age

Results by sex and age are illustrated in Figure 2 and Tables S6–S9. PM_2.5_ exposure was associated with an increase in overall ambulance attendances for women but not men, whereas injury-related ambulance attendances were elevated for both. Several health service use reductions were detectable among men only, including cardiovascular-related hospital admissions and emergency presentations, and mental health hospital admissions. Other detectable effect differences between men and women were too inconsistent across time stratifications to draw conclusions. Similarly, there were few notable differences in PM_2.5_ effects on healthcare use between older and younger cohort members.

**Figure 2.**
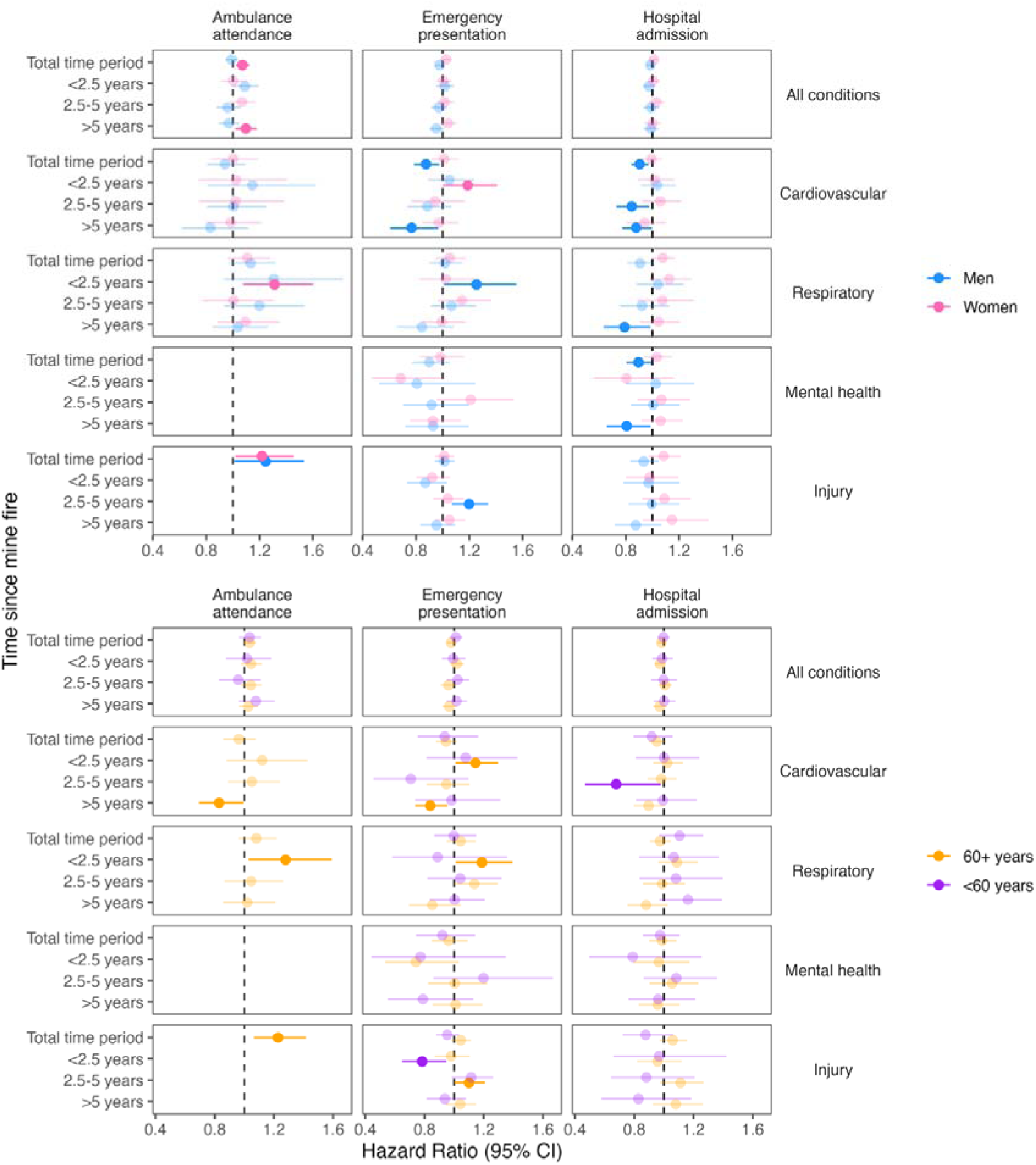
Changes in hospital admissions, emergency presentations, and ambulance attendances in association with a 10µg/m^3^ increase in daily mean mine fire-related PM_2.5_ exposure, by sex and age group

## 4 Discussion

This analysis examined the impact of PM_2.5_ exposure from the 2014 Hazelwood coal mine fire on ambulance attendances, emergency presentations, and hospital admissions over the following eight years with a focus on variation by time, condition, sex, and age group. We found no evidence that mine fire-related PM_2.5_ affected overall health service use, whether across the full study period or stratified by time. However, there was some evidence that PM_2.5_ exposure was associated with increased health service use for respiratory and cardiovascular conditions in the first 2.5 years following exposure, with effects which attenuated over time.

To our knowledge, there are no studies of long-term effects of smoke exposure from coal mine fires, or even withing the broader category of landscape fires, on health service use outside of the Hazelwood Health Study (13). The present findings are broadly consistent with other Hazelwood Health Study analyses using surveys, clinical follow-ups, and identified data linkage studies. For example, analyses of the Adult Survey cohort suggested that while smoke exposure from the mine fire initially worsened cardiovascular and respiratory health, these effects may have resolved over time (19,29).

However, the present findings contrast with results from Hazelwood Health Study analyses of anonymised, population-level health service use data, which found stronger evidence that exposure to PM_2.5_ from the mine fire resulted in a sustained increase in health service use. These included elevated risks of short-term hospital admissions and emergency presentations (14), ambulance attendances (15), mental health service use (33), and primary care and pharmaceutical service utilisation (34,35), particularly for respiratory and cardiovascular-related conditions. There was also evidence of a long-term increase in overall hospital admissions and emergency presentations in the eight years following the mine fire (16). The divergence between findings from these anonymised, population-level studies and the current individual-level analysis may be due in part to immortal time bias, or selection of study participants after an exposure had time to affect mortality (36). The exposed cohort can be depleted of the most vulnerable individuals, meaning the surviving members represent the healthiest members of the whole cohort. This can either falsely reduce the statistical association between an exposure and outcome, or even make it appear protective. In contrast, population-level, routinely-collected administrative data collects information on individuals before and after an exposure, therefore capturing changes due to effects on mortality.

There are several reasons to think immortal time bias affected the Hazelwood Health Study Adult Survey cohort. For instance, analysis of anonymous, population-level mortality data found an increase in cardiovascular-related deaths in Morwell during the six months following the fire (37). Notably, the increase in cardiovascular death after the fire was apparent in men, indicating this group would be most affected by immortal time bias; accordingly, the current analysis found PM_2.5_ exposure was associated with reduced cardiovascular hospital admissions and emergency presentations among men who survived long enough to join the cohort. This likely biased Adult Survey cohort members towards healthier survivors, which suggests that the associations observed in this study were an underestimate of the mine fire’s actual impacts on community health and healthcare burden. However, there are other important differences in study design – sample size, follow-up period, exposure resolution, availability of individual-level exposure and confounding variables – that contribute to divergent findings.

Within this context, we identified two associations that were more likely to reflect real harms from exposure to fire-related PM_2.5_. The first was the increase in respiratory-related ambulance attendances and possibly hospital admissions and emergency presentations, which attenuated over time. Perhaps the most consistently detected effect of the Hazelwood Health Study is that mine fire harmed respiratory health, according to analysis of both the Adult Survey cohort (14,15,19,28,38–42) and anonymous population-level administrative data (16,34,35), and that these effects attenuated over time, indicating potential recovery (29).

The second association that could indicate real harm from exposure to fire-related PM_2.5_ was the increase in injury-related ambulance attendances. Though an effect on injuries has not been consistently detected (15,19,20), analysis of anonymous health and mortality records found areas most affected by smoke had an increase in injury-related deaths during and shortly after the mine fire (37), and in the 8 years post-fire, there were substantial increases in injury-related hospital admittances and emergency presentations (16).

Elsewhere, ambient PM_2.5_ has been found to be associated with risk of fall-related injuries (43). That this effect was detectable in the current analysis despite immortal time bias – including people who died of injuries shortly after the fire and before they could participate in the Adult Survey cohort (37) – suggests the true effect is even larger. While the mechanism is not yet clear, one possible explanation relates to neurological effects of exposure to fire-related PM_2.5_ (37). For instance, specific components of mine fire smoke like Polycyclic Aromatic Hydrocarbons (PAHs) may accelerate neurodegeneration (44,45).

### 4.1 Strengths and limitations

This analysis had numerous strengths, including linkage between a cohort survey containing detailed individual-level confounder data with four separate administrative healthcare datasets, a relatively long follow-up period, and the ability to compare the current findings with other studies of the same phenomenon with different analytical approaches. In addition, analyses used individual-level measures of fire-related PM_2.5_.

The major limitation is the participation bias as well as selection bias due to effects of the mine fire on mortality in the months during and after it was extinguished (37). The modest sample size limited statistical power and made analysing some condition subcategories unfeasible. The outcome, healthcare utilisation, is not a direct measure of health outcomes and can be influenced by other factors such as socioeconomic status, which may have a bi-directional relationship with both exposure and outcome. There are differences in both coding standards and accuracy of diagnosed conditions across health services; this heterogeneity may limit comparability of condition effects across services.

## 5 Conclusions

In line with previous findings, we found evidence that smoke exposure from the Hazelwood coal mine fire worsened respiratory health and increased the long-term likelihood of injuries, as measured by health service use up to 8 years following exposure. The latter effect in particular raised questions about more complex mechanisms of harms from the mine fire. Immortal time bias due to the time elapsed between the fire and recruitment likely means the observed effects were a substantial underestimate.

## 6 Supplementary materials

### 6.1 Funding

This study was funded by the Department of Health, State Government of Victoria, though it represents the views of the authors and not the Department.

### 6.2 Supplementary tables

**Table S1.** Categories of condition types collected from hospital admissions, emergency department presentations, and ambulance attendances.

| Conditions | Hospital admissions/ Emergency presentations | Ambulance attendances |
| --- | --- | --- |
| Cardiovascular | I00-I99, G45, G46 | Acute coronary syndrome, acute myocardial infarction, angina, stroke, transient ischaemic attack, intracranial haemorrhage, cardiac arrest, cardiac failure, arrhythmia, aortic dissection, deep vein thrombosis, pulmonary embolism, acute pulmonary oedema |
| Respiratory | J00-J99 | Asthma, chronic obstructive pulmonary disease, cough, bronchitis, pneumonia, bronchiolitis, pulmonary aspiration, flu-like illness, pneumothorax, respiratory failure, respiratory arrest, respiratory tract infection, croup, chest infection, short of breath |
| Mental health | F00-F99 | Anxiety, panic attack, depression, emotional distress, psychiatric episode, eating disorder, sleep disorder, self-harm ideation, suicidal ideation |
| Injuries | S00-S99 | Laceration, fracture/s, wound/ puncture, burn/s, head injury |

**Table S2.** Changes in hospital admissions, emergency presentations, and ambulance attendances in association with a 10µg/m^3^ increase in daily mean mine fire-related PM_2.5_ exposure using the primary diagnosis code for emergency presentations and hospital admissions (underlying data for Figure 1)

|  | Ambulance attendances |  | Emergency presentations |  | Hospital admissions |  |
| --- | --- | --- | --- | --- | --- | --- |
|  | Hazard Ratio (95% CI) | p-value | Hazard Ratio (95% CI) | p-value | Hazard Ratio (95% CI) | p-value |
| **All conditions** |  |  |  |  |  |  |
| Total time period | 1.03 (0.99, 1.07) | 0.127 | 1.00 (0.98, 1.03) | 0.966 | 1.00 (0.97, 1.02) | 0.692 |
| <2.5 years | 1.04 (0.97, 1.11) | 0.244 | 1.01 (0.97, 1.06) | 0.597 | 0.99 (0.95, 1.02) | 0.426 |
| 2.5-5 years | 1.01 (0.95, 1.07) | 0.780 | 0.99 (0.95, 1.04) | 0.825 | 1.01 (0.97, 1.05) | 0.717 |
| >5 years | 1.03 (0.98, 1.09) | 0.241 | 1.00 (0.96, 1.04) | 0.899 | 0.99 (0.95, 1.04) | 0.702 |
| **Cardiovascular** |  |  |  |  |  |  |
| Total time period | 0.96 (0.86, 1.07) | 0.455 | 0.95 (0.88, 1.02) | 0.155 | **0.94 (0.89, 1.00)** | **0.036** |
| <2.5 years | 1.10 (0.88, 1.37) | 0.390 | **1.13 (1.01, 1.26)** | **0.041** | 1.02 (0.93, 1.13) | 0.675 |
| 2.5-5 years | 0.99 (0.83, 1.18) | 0.936 | 0.91 (0.79, 1.06) | 0.222 | 0.94 (0.85, 1.04) | 0.227 |
| >5 years | 0.89 (0.74, 1.06) | 0.193 | **0.88 (0.78, 0.99)** | **0.037** | 0.90 (0.82, 1.00) | 0.054 |
| **Respiratory** |  |  |  |  |  |  |
| Total time period | 1.11 (1.00, 1.24) | 0.060 | 1.04 (0.96, 1.13) | 0.342 | 1.01 (0.95, 1.07) | 0.806 |
| <2.5 years | **1.31 (1.04, 1.64)** | **0.020** | 1.12 (0.95, 1.33) | 0.171 | 1.09 (0.97, 1.22) | 0.135 |
| 2.5-5 years | 1.08 (0.90, 1.29) | 0.412 | 1.11 (0.98, 1.26) | 0.087 | 1.03 (0.90, 1.17) | 0.712 |
| >5 years | 1.06 (0.92, 1.22) | 0.449 | 0.94 (0.82, 1.08) | 0.363 | 0.94 (0.83, 1.06) | 0.295 |
| **Mental health** |  |  |  |  |  |  |
| Total time period | 0.92 (0.73, 1.16) | 0.480 | 0.95 (0.84, 1.08) | 0.433 | 0.99 (0.92, 1.07) | 0.866 |
| <2.5 years |  |  | 0.76 (0.56, 1.02) | 0.071 | 0.89 (0.72, 1.09) | 0.246 |
| 2.5-5 years |  |  | 1.11 (0.91, 1.36) | 0.306 | 1.07 (0.95, 1.21) | 0.243 |
| >5 years |  |  | 0.93 (0.79, 1.11) | 0.427 | 0.97 (0.86, 1.10) | 0.669 |
| **Injury** |  |  |  |  |  |  |
| Total time period | **1.23 (1.08, 1.40)** | **0.003** | 1.01 (0.96, 1.06) | 0.674 | 1.02 (0.94, 1.11) | 0.638 |
| <2.5 years | 1.25 (0.87, 1.81) | 0.238 | **0.89 (0.79, 0.99)** | **0.038** | 0.98 (0.85, 1.14) | 0.838 |
| 2.5-5 years | **1.21 (1.00, 1.46)** | **0.049** | **1.13 (1.04, 1.22)** | **0.003** | 1.06 (0.93, 1.20) | 0.375 |
| >5 years | 1.24 (1.00, 1.54) | 0.053 | 1.00 (0.92, 1.09) | 0.967 | 1.01 (0.87, 1.18) | 0.911 |

**Table S3.** Changes in hospital admissions, emergency presentations, and ambulance attendances in association with a 10µg/m^3^ increase in daily mean mine fire-related PM_2.5_ exposure, sensitivity analysis including both Morwell and Sale participants using the primary diagnosis code for emergency presentations and hospital admissions

|  | Ambulance attendances |  | Emergency presentations |  | Hospital admissions |  |
| --- | --- | --- | --- | --- | --- | --- |
|  | Hazard Ratio (95% CI) | p-value | Hazard Ratio (95% CI) | p-value | Hazard Ratio (95% CI) | p-value |
| **All conditions** |  |  |  |  |  |  |
| Total time period | 1.03 (0.99, 1.06) | 0.166 | 1.00 (0.97, 1.02) | 0.882 | 0.99 (0.97, 1.02) | 0.675 |
| <2.5 years | **1.08 (1.02, 1.15)** | **0.007** | 0.99 (0.95, 1.03) | 0.713 | 0.99 (0.95, 1.02) | 0.387 |
| 2.5-5 years | 1.01 (0.96, 1.07) | 0.663 | 0.99 (0.95, 1.04) | 0.776 | 1.01 (0.97, 1.05) | 0.683 |
| >5 years | 1.01 (0.96, 1.07) | 0.659 | 1.00 (0.97, 1.04) | 0.806 | 0.99 (0.95, 1.03) | 0.656 |
| **Cardiovascular** |  |  |  |  |  |  |
| Total time period | 0.96 (0.86, 1.08) | 0.493 | 0.95 (0.88, 1.02) | 0.154 | **0.95 (0.89, 1.00)** | **0.049** |
| <2.5 years | 1.15 (0.94, 1.40) | 0.180 | 1.08 (0.97, 1.20) | 0.156 | 0.99 (0.91, 1.09) | 0.909 |
| 2.5-5 years | 0.99 (0.83, 1.17) | 0.860 | 0.91 (0.80, 1.04) | 0.164 | 0.93 (0.84, 1.02) | 0.110 |
| >5 years | 0.88 (0.74, 1.05) | 0.164 | 0.90 (0.80, 1.00) | 0.061 | 0.93 (0.85, 1.02) | 0.132 |
| **Respiratory** |  |  |  |  |  |  |
| Total time period | 1.10 (0.99, 1.23) | 0.077 | 1.03 (0.95, 1.12) | 0.438 | 1.01 (0.95, 1.08) | 0.705 |
| <2.5 years | 1.21 (0.97, 1.50) | 0.101 | 1.01 (0.86, 1.19) | 0.865 | 1.09 (0.98, 1.21) | 0.127 |
| 2.5-5 years | 1.13 (0.95, 1.35) | 0.168 | 1.11 (0.99, 1.24) | 0.080 | 1.04 (0.93, 1.17) | 0.464 |
| >5 years | 1.05 (0.91, 1.21) | 0.497 | 0.99 (0.87, 1.11) | 0.811 | 0.93 (0.83, 1.04) | 0.228 |
| **Mental health** |  |  |  |  |  |  |
| Total time period | 0.91 (0.72, 1.15) | 0.430 | 0.96 (0.85, 1.08) | 0.476 | 0.97 (0.90, 1.05) | 0.524 |
| <2.5 years |  |  | 0.81 (0.64, 1.04) | 0.105 | 0.91 (0.78, 1.07) | 0.259 |
| 2.5-5 years |  |  | 1.05 (0.86, 1.29) | 0.637 | 1.05 (0.94, 1.18) | 0.378 |
| >5 years |  |  | 0.96 (0.82, 1.12) | 0.569 | 0.95 (0.84, 1.07) | 0.368 |
| **Injury** |  |  |  |  |  |  |
| Total time period | **1.22 (1.07, 1.39)** | **0.003** | 1.01 (0.96, 1.06) | 0.767 | 1.01 (0.93, 1.10) | 0.742 |
| <2.5 years | 1.09 (0.77, 1.54) | 0.636 | **0.90 (0.82, 0.99)** | **0.039** | 1.02 (0.89, 1.17) | 0.748 |
| 2.5-5 years | **1.25 (1.05, 1.48)** | **0.013** | **1.08 (1.00, 1.17)** | **0.038** | 1.04 (0.92, 1.18) | 0.555 |
| >5 years | **1.25 (1.04, 1.51)** | **0.020** | 1.02 (0.94, 1.10) | 0.683 | 0.99 (0.86, 1.13) | 0.859 |

Hazard Ratios (HRs) estimated from imputed PWP-CP model for both Morwell and Sale participants adjusting for confounding factors. Cause-specific hospitalisations and emergency presentations were obtained from all primary diagnoses.

**Table S4.** Changes in hospital admissions, emergency presentations, and ambulance attendances in association with a 10µg/m^3^ increase in daily mean mine fire-related PM_2.5_ exposure, sensitivity analyses using the principal diagnosis code for emergency presentations and hospital admissions

|  | Ambulance attendances |  | Emergency presentations |  | Hospital admissions |  |
| --- | --- | --- | --- | --- | --- | --- |
|  | Hazard Ratio (95% CI) | p-value | Hazard Ratio (95% CI) | p-value | Hazard Ratio (95% CI) | p-value |
| **All conditions** |  |  |  |  |  |  |
| Total time period | 1.03 (0.99, 1.07) | 0.113 | 1.00 (0.98, 1.03) | 0.893 | 1.00 (0.97, 1.02) | 0.751 |
| <2.5 years | 1.05 (0.98, 1.11) | 0.171 | 1.02 (0.97, 1.06) | 0.476 | 0.99 (0.95, 1.02) | 0.432 |
| 2.5-5 years | 1.01 (0.95, 1.07) | 0.785 | 0.99 (0.95, 1.04) | 0.689 | 1.01 (0.96, 1.05) | 0.789 |
| >5 years | 1.04 (0.98, 1.10) | 0.240 | 1.00 (0.96, 1.04) | 0.986 | 1.00 (0.95, 1.04) | 0.851 |
| **Cardiovascular** |  |  |  |  |  |  |
| Total time period | 0.97 (0.87, 1.09) | 0.647 | 0.95 (0.88, 1.02) | 0.176 | **0.90 (0.84, 0.96)** | **0.001** |
| <2.5 years | 1.10 (0.88, 1.37) | 0.388 | 1.11 (0.99, 1.25) | 0.075 | 1.00 (0.90, 1.11) | 0.979 |
| 2.5-5 years | 1.02 (0.87, 1.20) | 0.803 | 0.89 (0.78, 1.02) | 0.095 | **0.87 (0.77, 1.00)** | **0.042** |
| >5 years | 0.90 (0.75, 1.08) | 0.246 | 0.90 (0.79, 1.02) | 0.098 | **0.85 (0.76, 0.95)** | **0.006** |
| **Respiratory** |  |  |  |  |  |  |
| Total time period | **1.11 (1.00, 1.24)** | **0.047** | 1.05 (0.96, 1.14) | 0.300 | 1.06 (0.98, 1.15) | 0.156 |
| <2.5 years | **1.31 (1.05, 1.64)** | **0.018** | 1.13 (0.96, 1.34) | 0.141 | 1.13 (0.99, 1.29) | 0.070 |
| 2.5-5 years | 1.10 (0.92, 1.31) | 0.310 | 1.10 (0.97, 1.25) | 0.126 | 1.07 (0.91, 1.25) | 0.421 |
| >5 years | 1.06 (0.91, 1.22) | 0.457 | 0.95 (0.83, 1.09) | 0.452 | 0.99 (0.86, 1.14) | 0.913 |
| **Mental health** |  |  |  |  |  |  |
| Total time period | 0.92 (0.73, 1.16) | 0.498 | 0.97 (0.86, 1.10) | 0.641 | 1.00 (0.89, 1.13) | 0.953 |
| <2.5 years |  |  | 0.75 (0.55, 1.03) | 0.075 | 0.77 (0.49, 1.21) | 0.258 |
| 2.5-5 years |  |  | 1.12 (0.91, 1.38) | 0.281 | **1.33 (1.02, 1.72)** | **0.033** |
| >5 years |  |  | 0.97 (0.83, 1.14) | 0.715 | 0.93 (0.77, 1.12) | 0.443 |
| **Injury** |  |  |  |  |  |  |
| Total time period | **1.22 (1.07, 1.40)** | **0.004** | 1.01 (0.96, 1.06) | 0.778 | 1.05 (0.96, 1.15) | 0.271 |
| <2.5 years | 1.27 (0.88, 1.82) | 0.204 | **0.89 (0.79, 0.99)** | **0.039** | 1.04 (0.88, 1.23) | 0.632 |
| 2.5-5 years | **1.22 (1.00, 1.48)** | **0.048** | **1.12 (1.03, 1.21)** | **0.008** | 1.06 (0.92, 1.23) | 0.422 |
| >5 years | 1.22 (0.98, 1.52) | 0.081 | 1.00 (0.92, 1.09) | 0.988 | 1.05 (0.90, 1.22) | 0.546 |

Hazard Ratios (HRs) estimated from imputed PWP-CP model for Morwell participants adjusting for confounding factors. Cause-specific hospitalisations and emergency presentations were obtained from all principal diagnoses.

**Table S5.** Changes in hospital admissions, emergency presentations, and ambulance attendances in association with a 10µg/m^3^ increase in daily mean mine fire-related PM_2.5_ exposure, sensitivity analysis with counts capped at 5 services using the primary diagnosis code for emergency presentations and hospital admissions

|  | Ambulance attendances |  | Emergency presentations |  | Hospital admissions |  |
| --- | --- | --- | --- | --- | --- | --- |
|  | Hazard Ratio (95% CI) | p-value | Hazard Ratio (95% CI) | p-value | Hazard Ratio (95% CI) | p-value |
| **All conditions** |  |  |  |  |  |  |
| Total time period | 1.04 (1.00, 1.08) | 0.051 | 1.00 (0.98, 1.03) | 0.754 | 1.00 (0.97, 1.03) | 0.944 |
| <2.5 years | 1.05 (0.98, 1.12) | 0.171 | 1.01 (0.97, 1.06) | 0.588 | 0.99 (0.95, 1.03) | 0.537 |
| 2.5-5 years | 1.01 (0.94, 1.08) | 0.885 | 1.01 (0.96, 1.06) | 0.675 | 1.02 (0.97, 1.07) | 0.405 |
| >5 years | 1.06 (0.99, 1.12) | 0.082 | 0.99 (0.94, 1.04) | 0.713 | 0.99 (0.94, 1.04) | 0.698 |
| **Cardiovascular** |  |  |  |  |  |  |
| Total time period | 0.96 (0.86, 1.07) | 0.455 | 0.95 (0.88, 1.02) | 0.155 | **0.94 (0.89, 1.00)** | **0.036** |
| <2.5 years | 1.10 (0.88, 1.37) | 0.390 | **1.13 (1.01, 1.26)** | **0.041** | 1.02 (0.93, 1.13) | 0.675 |
| 2.5-5 years | 0.99 (0.83, 1.18) | 0.936 | 0.91 (0.79, 1.06) | 0.222 | 0.94 (0.85, 1.04) | 0.227 |
| >5 years | 0.89 (0.74, 1.06) | 0.193 | **0.88 (0.78, 0.99)** | **0.037** | 0.90 (0.82, 1.00) | 0.054 |
| **Respiratory** |  |  |  |  |  |  |
| Total time period | 1.11 (1.00, 1.24) | 0.060 | 1.04 (0.96, 1.13) | 0.342 | 1.01 (0.95, 1.07) | 0.806 |
| <2.5 years | **1.31 (1.04, 1.64)** | **0.020** | 1.12 (0.95, 1.33) | 0.171 | 1.09 (0.97, 1.22) | 0.135 |
| 2.5-5 years | 1.08 (0.90, 1.29) | 0.412 | 1.11 (0.98, 1.26) | 0.087 | 1.03 (0.90, 1.17) | 0.712 |
| >5 years | 1.06 (0.92, 1.22) | 0.449 | 0.94 (0.82, 1.08) | 0.363 | 0.94 (0.83, 1.06) | 0.295 |
| **Mental health** |  |  |  |  |  |  |
| Total time period | 0.92 (0.73, 1.16) | 0.480 | 0.95 (0.84, 1.08) | 0.433 | 0.99 (0.92, 1.07) | 0.866 |
| <2.5 years |  |  | 0.76 (0.56, 1.02) | 0.071 | 0.89 (0.72, 1.09) | 0.246 |
| 2.5-5 years |  |  | 1.11 (0.91, 1.36) | 0.306 | 1.07 (0.95, 1.21) | 0.243 |
| >5 years |  |  | 0.93 (0.79, 1.11) | 0.427 | 0.97 (0.86, 1.10) | 0.669 |
| **Injury** |  |  |  |  |  |  |
| Total time period | **1.23 (1.08, 1.40)** | **0.003** | 1.01 (0.96, 1.06) | 0.674 | 1.02 (0.94, 1.11) | 0.638 |
| <2.5 years | 1.25 (0.87, 1.81) | 0.238 | **0.89 (0.79, 0.99)** | **0.038** | 0.98 (0.85, 1.14) | 0.838 |
| 2.5-5 years | **1.21 (1.00, 1.46)** | **0.049** | **1.13 (1.04, 1.22)** | **0.003** | 1.06 (0.93, 1.20) | 0.375 |
| >5 years | 1.24 (1.00, 1.54) | 0.053 | 1.00 (0.92, 1.09) | 0.967 | 1.01 (0.87, 1.18) | 0.911 |

**Table S6.**
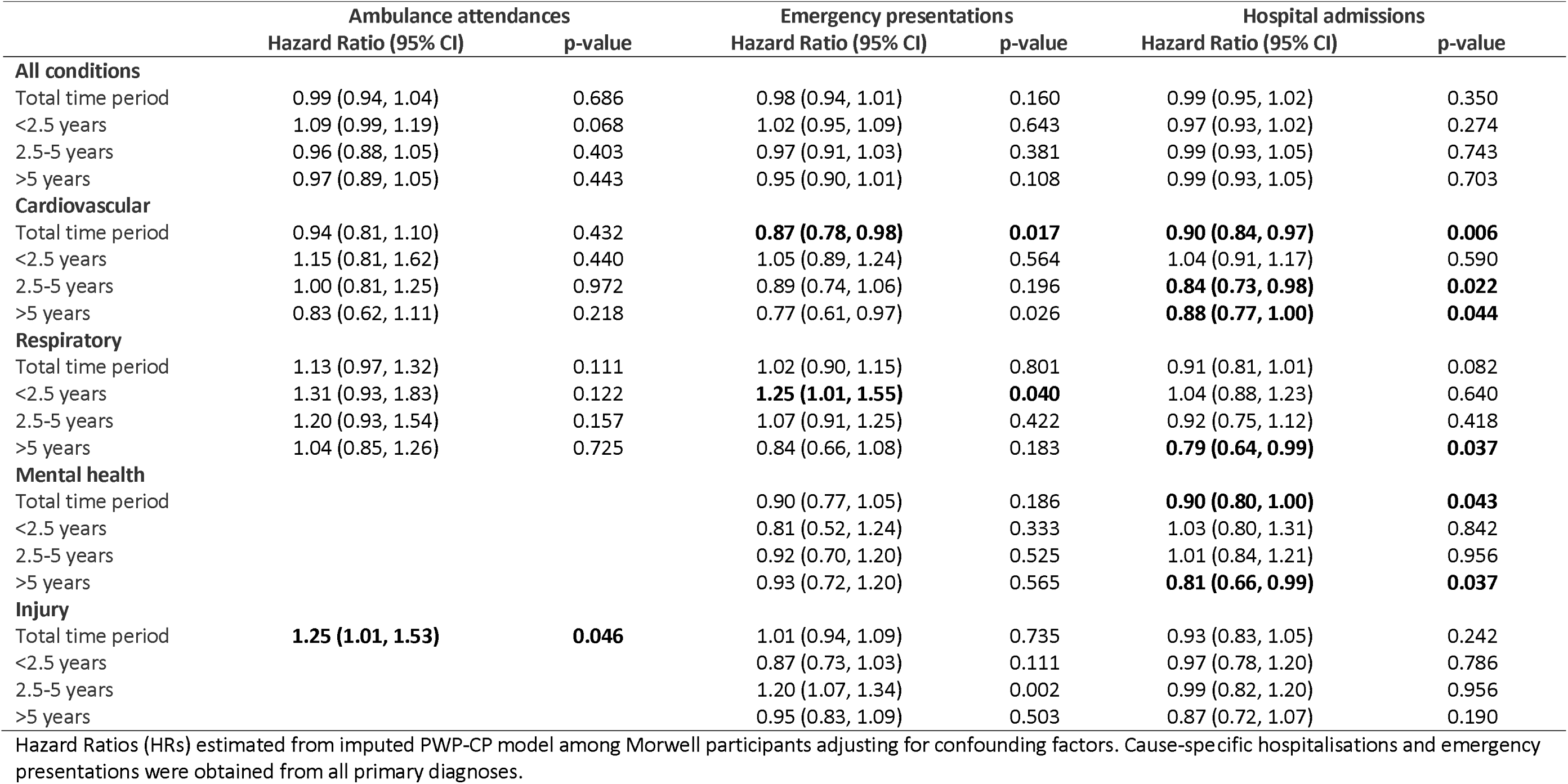
Changes in hospital admissions, emergency presentations, and ambulance attendances in association with a 10µg/m^3^ increase in daily mean mine fire-related PM_2.5_ exposure for male participants using the primary diagnosis code for emergency presentations and hospital admissions

|  | Ambulance attendances |  | Emergency presentations |  | Hospital admissions |  |
| --- | --- | --- | --- | --- | --- | --- |
|  | Hazard Ratio (95% CI) | p-value | Hazard Ratio (95% CI) | p-value | Hazard Ratio (95% CI) | p-value |
| **All conditions** |  |  |  |  |  |  |
| Total time period | 0.99 (0.94, 1.04) | 0.686 | 0.98 (0.94, 1.01) | 0.160 | 0.99 (0.95, 1.02) | 0.350 |
| <2.5 years | 1.09 (0.99, 1.19) | 0.068 | 1.02 (0.95, 1.09) | 0.643 | 0.97 (0.93, 1.02) | 0.274 |
| 2.5-5 years | 0.96 (0.88, 1.05) | 0.403 | 0.97 (0.91, 1.03) | 0.381 | 0.99 (0.93, 1.05) | 0.743 |
| >5 years | 0.97 (0.89, 1.05) | 0.443 | 0.95 (0.90, 1.01) | 0.108 | 0.99 (0.93, 1.05) | 0.703 |
| **Cardiovascular** |  |  |  |  |  |  |
| Total time period | 0.94 (0.81, 1.10) | 0.432 | **0.87 (0.78, 0.98)** | **0.017** | **0.90 (0.84, 0.97)** | **0.006** |
| <2.5 years | 1.15 (0.81, 1.62) | 0.440 | 1.05 (0.89, 1.24) | 0.564 | 1.04 (0.91, 1.17) | 0.590 |
| 2.5-5 years | 1.00 (0.81, 1.25) | 0.972 | 0.89 (0.74, 1.06) | 0.196 | **0.84 (0.73, 0.98)** | **0.022** |
| >5 years | 0.83 (0.62, 1.11) | 0.218 | 0.77 (0.61, 0.97) | 0.026 | **0.88 (0.77, 1.00)** | **0.044** |
| **Respiratory** |  |  |  |  |  |  |
| Total time period | 1.13 (0.97, 1.32) | 0.111 | 1.02 (0.90, 1.15) | 0.801 | 0.91 (0.81, 1.01) | 0.082 |
| <2.5 years | 1.31 (0.93, 1.83) | 0.122 | **1.25 (1.01, 1.55)** | **0.040** | 1.04 (0.88, 1.23) | 0.640 |
| 2.5-5 years | 1.20 (0.93, 1.54) | 0.157 | 1.07 (0.91, 1.25) | 0.422 | 0.92 (0.75, 1.12) | 0.418 |
| >5 years | 1.04 (0.85, 1.26) | 0.725 | 0.84 (0.66, 1.08) | 0.183 | **0.79 (0.64, 0.99)** | **0.037** |
| **Mental health** |  |  |  |  |  |  |
| Total time period |  |  | 0.90 (0.77, 1.05) | 0.186 | **0.90 (0.80, 1.00)** | **0.043** |
| <2.5 years |  |  | 0.81 (0.52, 1.24) | 0.333 | 1.03 (0.80, 1.31) | 0.842 |
| 2.5-5 years |  |  | 0.92 (0.70, 1.20) | 0.525 | 1.01 (0.84, 1.21) | 0.956 |
| >5 years |  |  | 0.93 (0.72, 1.20) | 0.565 | **0.81 (0.66, 0.99)** | **0.037** |
| **Injury** |  |  |  |  |  |  |
| Total time period | **1.25 (1.01, 1.53)** | **0.046** | 1.01 (0.94, 1.09) | 0.735 | 0.93 (0.83, 1.05) | 0.242 |
| <2.5 years |  |  | 0.87 (0.73, 1.03) | 0.111 | 0.97 (0.78, 1.20) | 0.786 |
| 2.5-5 years |  |  | 1.20 (1.07, 1.34) | 0.002 | 0.99 (0.82, 1.20) | 0.956 |
| >5 years |  |  | 0.95 (0.83, 1.09) | 0.503 | 0.87 (0.72, 1.07) | 0.190 |

Hazard Ratios (HRs) estimated from imputed PWP-CP model among Morwell participants adjusting for confounding factors. Cause-specific hospitalisations and emergency presentations were obtained from all primary diagnoses.

**Table S7.** Changes in hospital admissions, emergency presentations, and ambulance attendances in association with a 10µg/m^3^ increase in daily mean mine fire-related PM_2.5_ exposure for female participants using the primary diagnosis code for emergency presentations and hospital admissions

|  | Ambulance attendances |  | Emergency presentations |  | Hospital admissions |  |
| --- | --- | --- | --- | --- | --- | --- |
|  | Hazard Ratio (95% CI) | p-value | Hazard Ratio (95% CI) | p-value | Hazard Ratio (95% CI) | p-value |
| **All conditions** |  |  |  |  |  |  |
| Total time period | **1.07 (1.02, 1.13)** | **0.006** | 1.02 (0.99, 1.06) | 0.189 | 1.01 (0.98, 1.04) | 0.530 |
| <2.5 years | 1.00 (0.91, 1.11) | 0.978 | 1.01 (0.95, 1.06) | 0.796 | 1.01 (0.96, 1.06) | 0.805 |
| 2.5-5 years | 1.07 (0.98, 1.17) | 0.141 | 1.02 (0.95, 1.08) | 0.629 | 1.03 (0.97, 1.09) | 0.330 |
| >5 years | **1.10 (1.02, 1.18)** | **0.014** | 1.04 (0.98, 1.10) | 0.184 | 1.00 (0.94, 1.06) | 0.999 |
| **Cardiovascular** |  |  |  |  |  |  |
| Total time period | 1.00 (0.84, 1.19) | 0.985 | 1.01 (0.92, 1.12) | 0.829 | 0.99 (0.92, 1.08) | 0.877 |
| <2.5 years | 1.02 (0.74, 1.41) | 0.892 | **1.19 (1.00, 1.41)** | **0.049** | 1.02 (0.89, 1.17) | 0.811 |
| 2.5-5 years | 1.02 (0.75, 1.39) | 0.906 | 0.94 (0.76, 1.16) | 0.586 | 1.06 (0.92, 1.22) | 0.411 |
| >5 years | 0.98 (0.79, 1.22) | 0.877 | 0.97 (0.85, 1.12) | 0.692 | 0.94 (0.81, 1.10) | 0.443 |
| **Respiratory** |  |  |  |  |  |  |
| Total time period | 1.11 (0.96, 1.28) | 0.167 | 1.05 (0.95, 1.17) | 0.351 | 1.08 (1.00, 1.17) | 0.065 |
| <2.5 years | **1.31 (1.08, 1.60)** | **0.009** | 1.02 (0.83, 1.27) | 0.833 | 1.12 (0.98, 1.29) | 0.105 |
| 2.5-5 years | 1.00 (0.77, 1.31) | 0.975 | 1.15 (0.96, 1.37) | 0.129 | 1.08 (0.89, 1.31) | 0.453 |
| >5 years | 1.09 (0.89, 1.35) | 0.402 | 0.99 (0.85, 1.17) | 0.944 | 1.05 (0.91, 1.21) | 0.534 |
| **Mental health** |  |  |  |  |  |  |
| Total time period |  |  | 0.98 (0.83, 1.16) | 0.842 | 1.03 (0.93, 1.14) | 0.529 |
| <2.5 years |  |  | 0.69 (0.47, 1.01) | 0.058 | 0.80 (0.56, 1.16) | 0.245 |
| 2.5-5 years |  |  | 1.21 (0.96, 1.53) | 0.116 | 1.07 (0.89, 1.28) | 0.487 |
| >5 years |  |  | 0.93 (0.76, 1.14) | 0.467 | 1.06 (0.92, 1.23) | 0.427 |
| **Injury** |  |  |  |  |  |  |
| Total time period | **1.22 (1.02, 1.46)** | **0.036** | 1.01 (0.94, 1.09) | 0.749 | 1.08 (0.97, 1.21) | 0.156 |
| <2.5 years |  |  | 0.92 (0.80, 1.06) | 0.236 | 0.98 (0.80, 1.20) | 0.830 |
| 2.5-5 years |  |  | 1.04 (0.93, 1.16) | 0.502 | 1.09 (0.92, 1.29) | 0.314 |
| >5 years |  |  | 1.05 (0.94, 1.17) | 0.375 | 1.15 (0.93, 1.42) | 0.209 |

Hazard Ratios (HRs) estimated from imputed PWP-CP model among Morwell participants adjusting for confounding factors. Cause-specific hospitalisations and emergency presentations were obtained from all primary diagnoses.

**Table S8.** Changes in hospital admissions, emergency presentations, and ambulance attendances in association with a 10µg/m^3^ increase in daily mean mine fire-related PM_2.5_ exposure for participants aged under 60 using the primary diagnosis code for emergency presentations and hospital admissions

|  | Ambulance attendances |  | Emergency presentations |  | Hospital admissions |  |
| --- | --- | --- | --- | --- | --- | --- |
|  | Hazard Ratio (95% CI) | p-value | Hazard Ratio (95% CI) | p-value | Hazard Ratio (95% CI) | p-value |
| **All conditions** |  |  |  |  |  |  |
| Total time period | 1.04 (0.96, 1.11) | 0.344 | 1.01 (0.97, 1.06) | 0.572 | 1.00 (0.96, 1.04) | 0.902 |
| <2.5 years | 1.02 (0.88, 1.18) | 0.818 | 0.99 (0.92, 1.08) | 0.878 | 0.99 (0.92, 1.06) | 0.764 |
| 2.5-5 years | 0.96 (0.83, 1.11) | 0.568 | 1.02 (0.95, 1.10) | 0.539 | 1.00 (0.92, 1.09) | 0.971 |
| >5 years | 1.08 (0.96, 1.21) | 0.196 | 1.02 (0.95, 1.09) | 0.646 | 1.00 (0.93, 1.08) | 0.965 |
| **Cardiovascular** |  |  |  |  |  |  |
| Total time period |  |  | 0.94 (0.75, 1.16) | 0.556 | 0.92 (0.79, 1.06) | 0.244 |
| <2.5 years |  |  | 1.08 (0.81, 1.43) | 0.603 | 1.00 (0.81, 1.24) | 0.991 |
| 2.5-5 years |  |  | 0.71 (0.46, 1.10) | 0.125 | **0.68 (0.47, 0.98)** | **0.040** |
| >5 years |  |  | 0.98 (0.74, 1.31) | 0.907 | 0.99 (0.81, 1.22) | 0.961 |
| **Respiratory** |  |  |  |  |  |  |
| Total time period |  |  | 1.00 (0.87, 1.15) | 0.979 | 1.11 (0.97, 1.26) | 0.143 |
| <2.5 years |  |  | 0.89 (0.58, 1.36) | 0.586 | 1.07 (0.83, 1.37) | 0.605 |
| 2.5-5 years |  |  | 1.04 (0.82, 1.32) | 0.742 | 1.08 (0.84, 1.40) | 0.551 |
| >5 years |  |  | 1.00 (0.83, 1.21) | 0.966 | 1.16 (0.97, 1.39) | 0.106 |
| **Mental health** |  |  |  |  |  |  |
| Total time period |  |  | 0.92 (0.74, 1.14) | 0.455 | 0.97 (0.86, 1.11) | 0.695 |
| <2.5 years |  |  | 0.77 (0.44, 1.35) | 0.367 | 0.79 (0.50, 1.26) | 0.320 |
| 2.5-5 years |  |  | 1.20 (0.86, 1.67) | 0.286 | 1.08 (0.86, 1.36) | 0.488 |
| >5 years |  |  | 0.79 (0.55, 1.13) | 0.198 | 0.96 (0.76, 1.21) | 0.739 |
| **Injury** |  |  |  |  |  |  |
| Total time period |  |  | 0.95 (0.88, 1.03) | 0.247 | 0.88 (0.72, 1.06) | 0.186 |
| <2.5 years |  |  | **0.78 (0.65, 0.95)** | **0.012** | 0.97 (0.66, 1.42) | 0.867 |
| 2.5-5 years |  |  | 1.12 (0.98, 1.26) | 0.088 | 0.88 (0.64, 1.21) | 0.435 |
| >5 years |  |  | 0.94 (0.81, 1.08) | 0.373 | 0.83 (0.58, 1.19) | 0.305 |

Hazard Ratios (HRs) estimated from imputed PWP-CP model among Morwell participants adjusting for confounding factors. Cause-specific hospitalisations and emergency presentations were obtained from all primary diagnoses.

**Table S9.** Changes in hospital admissions, emergency presentations, and ambulance attendances in association with a 10µg/m^3^ increase in daily mean mine fire-related PM_2.5_ exposure for participants aged 60 or over

|  | Ambulance attendances |  | Emergency presentations |  | Hospital admissions |  |
| --- | --- | --- | --- | --- | --- | --- |
|  | Hazard Ratio (95% CI) | p-value | Hazard Ratio (95% CI) | p-value | Hazard Ratio (95% CI) | p-value |
| **All conditions** |  |  |  |  |  |  |
| Total time period | 1.03 (0.99, 1.08) | 0.099 | 0.98 (0.95, 1.01) | 0.150 | 0.98 (0.96, 1.01) | 0.246 |
| <2.5 years | 1.05 (0.98, 1.12) | 0.198 | 1.02 (0.97, 1.07) | 0.451 | 0.97 (0.94, 1.01) | 0.168 |
| 2.5-5 years | 1.04 (0.98, 1.12) | 0.216 | 0.96 (0.91, 1.02) | 0.183 | 1.01 (0.96, 1.05) | 0.746 |
| >5 years | 1.03 (0.96, 1.09) | 0.413 | 0.97 (0.92, 1.02) | 0.196 | 0.97 (0.93, 1.02) | 0.266 |
| **Cardiovascular** |  |  |  |  |  |  |
| Total time period | 0.96 (0.86, 1.08) | 0.512 | 0.95 (0.88, 1.02) | 0.142 | 0.95 (0.90, 1.01) | 0.112 |
| <2.5 years | 1.12 (0.88, 1.43) | 0.357 | **1.14 (1.01, 1.30)** | **0.034** | 1.02 (0.93, 1.13) | 0.644 |
| 2.5-5 years | 1.05 (0.89, 1.24) | 0.560 | 0.95 (0.81, 1.10) | 0.493 | 0.98 (0.89, 1.09) | 0.732 |
| >5 years | **0.83 (0.69, 0.99)** | **0.043** | **0.84 (0.74, 0.95)** | **0.008** | 0.90 (0.80, 1.01) | 0.067 |
| **Respiratory** |  |  |  |  |  |  |
| Total time period | 1.08 (0.96, 1.22) | 0.198 | 1.04 (0.95, 1.15) | 0.376 | 0.97 (0.91, 1.05) | 0.478 |
| <2.5 years | **1.28 (1.03, 1.59)** | **0.028** | **1.19 (1.01, 1.39)** | **0.037** | 1.09 (0.96, 1.23) | 0.176 |
| 2.5-5 years | 1.05 (0.87, 1.26) | 0.642 | 1.14 (1.00, 1.29) | 0.053 | 0.99 (0.86, 1.14) | 0.891 |
| >5 years | 1.02 (0.86, 1.21) | 0.827 | 0.85 (0.70, 1.05) | 0.126 | 0.88 (0.75, 1.03) | 0.105 |
| **Mental health** |  |  |  |  |  |  |
| Total time period |  |  | 0.96 (0.85, 1.09) | 0.552 | 0.99 (0.90, 1.09) | 0.811 |
| <2.5 years |  |  | 0.74 (0.53, 1.03) | 0.078 | 0.97 (0.79, 1.17) | 0.727 |
| 2.5-5 years |  |  | 1.00 (0.82, 1.22) | 0.964 | 1.06 (0.90, 1.23) | 0.496 |
| >5 years |  |  | 1.01 (0.86, 1.19) | 0.907 | 0.96 (0.83, 1.11) | 0.568 |
| **Injury** |  |  |  |  |  |  |
| Total time period | **1.23 (1.06, 1.42)** | **0.007** | 1.04 (0.98, 1.11) | 0.182 | 1.06 (0.97, 1.16) | 0.186 |
| <2.5 years |  |  | 0.98 (0.87, 1.11) | 0.746 | 0.96 (0.82, 1.12) | 0.589 |
| 2.5-5 years |  |  | **1.10 (1.00, 1.21)** | **0.046** | 1.11 (0.98, 1.27) | 0.114 |
| >5 years |  |  | 1.04 (0.94, 1.15) | 0.419 | 1.08 (0.92, 1.26) | 0.332 |

Hazard Ratios (HRs) estimated from imputed PWP-CP model among Morwell participants adjusting for confounding factors. Cause-specific hospitalisations and emergency presentations were obtained from all primary diagnoses.

## Data Availability

All data used in this study are confidential and cannot be shared.

